# Emergence and antibody evasion of BQ, BA.2.75 and SARS-CoV-2 recombinant sublineages in the face of maturing antibody breadth at the population level

**DOI:** 10.1101/2022.12.06.22283000

**Authors:** Anouschka Akerman, Vanessa Milogiannakis, Tyra Jean, Camille Esneau, Mariana Ruiz Silva, Timothy Ison, Christina Fitcher, Joseph A Lopez, Deborah Chandra, Zin Naing, Joanna Caguicla, Daiyang Li, Gregory Walker, Supavadee Amatayakul-Chantler, Nathan Roth, Sandro Manni, Thomas Hauser, Thomas Barnes, Anna Condylios, Malinna Yeang, Maureen Wong, Charles S.P. Foster, Kenta Sato, Sharon Lee, Yang Song, Lijun Mao, Allison Sigmund, Amy Phu, Ann Marie Vande More, Stephanie Hunt, Mark Douglas, Ian Caterson, Kerrie Sandgren, Rowena Bull, Andrew Lloyd, Jamie Triccas, Stuart Tangye, Nathan W Bartlett, David Darley, Gail Matthews, Damien J. Stark, William D. Rawlinson, Ben Murrell, Fabienne Brilot, Anthony L Cunningham, Anthony D. Kelleher, Anupriya Aggarwal, Stuart G. Turville

**Affiliations:** The Kirby Institute, University of New South Wales, New South Wales, Australia; Serology and Virology Division (SAViD), NSW Health Pathology, Randwick, Australia; Hunter Medical Research Institute, University of Newcastle; Brain Autoimmunity Group, Kids Neuroscience Centre, The Children’s Hospital at Westmead, Faculty of Medicine and Health, School of Medical Sciences, Sydney University of Sydney, Sydney Institute for Infectious Diseases, Sydney, New South WalesLambton Heights, Australia; Department of Bioanalytical Sciences, Plasma Product Development, Research & Development, CSL Behring, Australia; Department of Bioanalytical Sciences, Plasma Product Development, Research & Development, CSL Behring AG, Bern, Switzerland; Plasma Product Development, Research & Development, CSL Behring AG, Bern, Switzerland; Research & Education Network, Westmead Hospital, WSLHD, NSW, Australia; Royal Prince Alfred Hospital, SLHD, NSW Australia; The Westmead Institute for Medical Research, Westmead, New South Wales, Australia, School of Medical Sciences, Faculty of Medicine and Health, University of Sydney, Sydney, NSW, AustraliaVincent’s; Sydney Institute for Infectious Diseases and the Charles Perkins Centre, The University of Sydney. Camperdown, New South Wales, Australia; Garvan Institute of Medical Research, Sydney, New South Wales, Australia; St Vincent’s Hospital, Sydney, New South Wales, Australia; Molecular Diagnostic Medicine Laboratory, Sydpath, St Vincent’s Hospital, Sydney, New South Wales, Australia; Department of Microbiology, Tumor and Cell Biology, Karolinska Institutet, 171 77 Stockholm

**Author notes:** Corresponding Author:* Stuart G. Turville, The Kirby Institute, UNSW Australia, Office 529 Level 5 Wallace Wurth Building, UNSW, Sydney NSW 2052. Equal contribution (Anouschka Akerman, Vanessa Milogiannakis). Equal senior author (Anupriya Aggarwal, Stuart G. Turville).

## Abstract

The Omicron era of the COVID-19 pandemic commenced at the beginning of 2022 and whilst it started with primarily BA.1, it was latter dominated by BA.2 and related sub-lineages. Over the course of 2022, we monitored the potency and breadth of antibody neutralization responses to many emerging variants at two levels: (i) we tracked over 420,000 U.S. plasma donors over time through various vaccine booster roll outs and Omicron waves using sequentially collected IgG pools; (ii) we mapped the antibody response in individuals using blood from strigently curated vaccine and convalescent cohorts. In pooled IgG samples, we observed the maturation of neutralization breadth to Omicron variants over time through continuing vaccine and infection waves. Importantly, in many cases we observed increased antibody breadth to variants that were yet to be in circulation. Determination of viral neutralization at the cohort level supported equivalent coverage across prior and emerging variants with emerging isolates BQ.1.1, XBB.1, BR.2.1 and XBF the most evasive. Further, these emerging variants were resistant to Evusheld, whilst neutralization resistance to Sotrovimab was restricted to BQ.1.1 and XBF. We conclude at this current point in time that dominant variants can evade antibodies at levels equivalent to their most evasive lineage counterparts but sustain an entry phenotype that continues to promote an additional outgrowth advantage. In Australia, BR2.1 and XBF share this phenotype and are dominating across NSW and Victoria.

**Research in context:** *Evidence before this study:* Up until the BA.5 wave in mid 2022, many global waves were seeded by dominant variants such as Delta, Omicron BA.1 and Omicron BA.2. Following resolution of the BA.5, was the emergence of a pool of BA.4/5 and BA.2.75 sub-lineages accumulating clusters of similar polymorphisms located with the Receptor Binding Domain (RBD) of the Spike glycoprotein. Although iterative changes in the Spike increased the ability of each variant to navigate existing neutralising antibodies, it was unclear if this alone was sufficient to provide an outgrowth advantage to any one variant to fuel major case waves in global communities with high vaccine uptake and/or infection.

*Added value of this study:* Prior studies on incoming variants in Australian quarantine, highlighted the potential for Australia to represent a unique mix of cocirculating variants. Following the resolution of the BA.5 Omicron wave, many globally circulating variants appeared early on and ranged from BA.2.75 lineages, recombinants XBB.1, and XBC.1 in addition to many BA.5 derived BQ.1 lineages. Two additional lineages, the recombinant XBF and the BA.2.75 derived BR.2.1 also appeared and were uniquely enriched in Australia. Using 14 primary clinical isolates covering a continuum of circulating variants in Australia, we resolved neutralisation responses of 110 donors stringently documented for their vaccine and infection status over time. In addition, we also tested the well clinical utilised clinical monoclonals Evusheld and Sotrovimab. In addition to tracking donors, we also tracked immunity at the population level, using pooled IgG samples over time. The latter samples were the sum of 420,000 US plasma donors covering time periods of high-booster uptake alongside and in addition to large case waves. Whilst the above resolved the impact of Spike changes in neutralisations, we also tested each variant with respect to the efficiency of TMPRSS2 use, as this significantly influences viral tropism across the respiratory tract.

*Implications of all the available evidence:* All variants analysed herein have undertaken a convergent trajectory in accumulating a similar cluster of Spike polymorphisms. Many variants, including BQ.1.1, XBB.1, XBF and BR.2.1 have accumulated key changes that now render neutralisation responses lower in all cohorts and are neutralisation resistant to Evusheld. Whilst sotrovimab retained neutralisation capacity of many variants, there was significant reduction for variants BQ.1.1 and XBF. Impact of Spike changes on TMPRSS2 use were mixed and only one variant, BQ.1.2, had equal to increased usage relative to its parent BA.5. Analysis of neutralisation at the population level over time revealed two key observations. Firstly, whilst variants converged and lowered neutralisation responses, this reduction was negated over time with increasing neutralisation breadth. Secondly, responses to a variant proceeded its appearance and global circulation. In conclusion, whilst many variants are appearing and iterative changes in the spike will challenge antibody responses, increasing breadth in the community over time has enabled sufficient coverage to presently emerging variants. Furthermore, with the exception of BQ.1.2, viral use of TMPRSS2 has not increased and as such viral tropism towards epithelial cells of the upper respiratory tract we predict will be maintained.

## Introduction

The beginning of the COVID-19 pandemic saw severe acute respiratory syndrome coronavirus 2 (SARS-CoV-2) variants driving several waves of disease transmission globally. The initial lack of vaccines and often immature/germ line convalescent antibody responses ^2-6^ promoted small iterative changes in the virus genome during various global waves of infection. Changes primarily within the Spike glycoprotein were key to viral spread. Whilst many of these changes appeared in the N-terminal domain and within the receptor binding domain, resulting in decreased antibody binding, others accumulated at and around the furin cleavage site which supported greater viral fitness and cellular entry through the serine protease TMPRSS2 ^7-13^. The Delta variant highlighted how changes in viral entry and fitness combined with changes that enabled antibody evasion could drive significant waves worldwide.

Fortunately, towards the end of 2021, many vaccines were mobilized worldwide that led to greater than 88% vaccine efficacy to the dominant Delta variant ^14^. The subsequent arrival of two Omicron lineages BA.1 and BA.2 represented a significant shift in the pandemic at two key levels. Firstly, both sub-variants appeared with Spike glycoproteins that enabled significant evasion from both vaccine and/or infection-induced antibodies. Secondly, unlike Delta, BA.1 and BA.2 shifted away from efficient usage of TMPRSS2, which favored replication in upper respiratory tract epithelial cells ^8,15-18^. With the resolution of both BA.1 and BA.2 waves globally, the BA.2 sub-lineage, BA.5, propelled subsequent waves. This sub-lineage was defined by key changes in the receptor binding domain resulting in antibody evasion and greater use of TMPRSS2 relative to its parent BA.2 ^1^. The BA.5 wave has since been replaced by BA.5 and BA.2.75 sub-lineages that have accumulated additional changes across their genomes, especially within the Spike glycoprotein.

To resolve the relative threat of circulating variants in real-time, we have tested variants prevalent within the community using two approches: (i) isolation and whole genome sequencing of SARS-CoV-2 grown from nasopharyngeal swabs and; (ii) phenotypic characterization of antibody evasion and modes of viral entry. The latter was examined using four distinct approaches. We first evaluated neutralizing antibody evasion at the population level with pooled plasma from over 420,000 U.S. donors. The second approach involved testing of curated cohorts based on vaccine- and infection-history. Thirdly, we determined the ability of currently approved monoclonal antibody therapeutics to neutralize the circulating variants. Finally, we observed the consequences of these cumulative changes in Spike on TMPRSS2 use, as this can significantly influence viral entry fitness and epithelial cell tropism throughout the respiratory tract.

Based on the above, we observe the maturation of the neutralization responses to target a greater diversity of Omicron variants. At the viral level, we observe the convergence of Spike polymorphisms across many emerging lineages to significantly decrease the potency and breadth of the neutralization responses in all cohorts. This translated to the loss of Evusheld’s ability to neutralize key emerging variants XBB.1, BR.2.1, BQ.1.1 and XBF. In contrast, Sotrovimab retained potent neutralization activity to all tested variants with the exception of BQ.1.1 and XBF. With respect to viral entry, dominant circulating BA.2.75 and BQ sub-lineages such as XBF, BR.2.1 and BQ.1.1 exhibited similar but reduced TMPRSS2 use relative to BA.5. We conclude that whilst the increasing breadth of antibody responses is increasing pressure on the current variant pool, continued variant surveillance will be important in determining the trajectory of current and future variant pools with respect to antibody evasion and tropism.

## Methods

### Human sera

The ADAPT cohort is composed of RT-PCR–confirmed convalescent individuals (including some subsequently vaccinated) recruited in Australia since 2020^2^. Donors within this cohort completed their first vaccine doses (ChAdOx1 or BNT162b2; See Supplementary Table S3) and booster doses (BNT162b2 or mRNA-1273) in January to February 2022. Female to male ratios were 0.5:1.0 for this cohort, with a donor median age of 51 years. The Australian Vaccine, Infection and Immunology Collaborative Research cohort (VIIM) cohort consists of serum samples from 75 participants collected between July 2022 – October 2022. Serum samples were collected at 12 months post second dose of Pfizer Comirnaty from two sites; Westmead Hospital and Royal Prince Alfred Hospital, NSW, Australia. Female to male ratio was 1.02: 1.0, with a donor median age of 50.6 years. Data were collected on SARS-CoV2 infections for all participants.

### Ethics statement

All human serum samples were obtained with written informed consent from the participants (2020/ETH00964; 2020/ETH02068; 2019/ETH03336; 2021/ETH00180; 2021/ETH0042). All primary isolates used herein were obtained from de-identified remnant diagnostic swabs that had completed all diagnostic testing under approval by the New South Wales Chief Health Officer following independent scientific review and as outlined in the ADAPT ethics protocol 2020/ETH00964.

### Other immunoglobulin products

Clinical grade Sotrovimab (62.5 mg/mL; NDC 0173-0901-86) was kindly provided by GSK Healthcare while clinical grade Cilgavimab and Tixagevimab (100 mg/mL each; AstraZeneca) were kindly provided by Dr Sarah Sasson (Kirby Institute, UNSW). Cilgavimab and Tixagevimab were mixed in equal volumes to generate the monoclonal antibody cocktail Evusheld. All monoclonal antibodies were tested at a starting concentration of 10 µg/mL and diluted three-fold in an eight step dilution series.

### Polyclonal immunoglobulin preparations and anti-SARs-CoV-2 hyperimmune globulin

The immunoglobulins used herein were purified using the licensed and fully validated immunoglobulin manufacturing process used for Privigen ^19^, notionally similar to others^3^. Thirty seven poly-Ig batches were manufactured using the Privigen process^19^ and included US plasma collected by plasmapheresis from a mixture of vaccinated (SARS-CoV-2 mRNA vaccines), convalescent and non-convalescent donors (source plasma, n >10,000 donors per batch) collected over the period of August 2021 to October 2022.

### Cell culture

HEK293T cells stably expressing human ACE2 and TMPRSS2 were generated by lentiviral transductions as previously described^2,17^. A highly permissive clone (HAT-24) was identified through clonal selection and used for this study. The HAT-24 line has been extensively cross-validated with the VeroE6 cell line^17^. VeroE6-TMPRSS2 (Vero-T) cells were kindly provided by Associate Professor Daniel Watterson (University of Queensland). HAT-24, VeroE6-TMPRSS2 cells were cultured in Dulbecco’s Modified Eagle Medium (Gibco, 11995073) containing 10% foetal bovine serum (Gibco, 10099141; DMEM-10%FBS) and VeroE6 cells (ATCC® CRL-1586™) in Minimal Essential Medium (Gibco, 11090099) containing 10% FBS and 1% penicillin-streptomycin (Gibco, 15140122; MEM-10%FBS). pBEC and alveolar epithelial cultures were grown and differentiated until confluent in complete Bronchial Epithelial Cell Growth Basal Medium (Lonza, CC-3171) before use for air-liquid interface experiments. All cells were incubated at 37°C, 5% CO_**2**_ and >90% relative humidity. For VeroE6-TMPRSS2 cell line authentication was performed as previously described^2,17^. The STR profiling of HAT-24 has been previously described^17^. All cell lines tested negative for mycoplasma.

### Visualizing variant dynamics

Variant competition was inferred using the global GISAID dataset (bulk .fasta download from 2022/12/2022), excluding any data older than 100 days. GISAID sequences were processed and assigned to clades via NextClade (using the BA.2 reference set, “sars-cov-2-21L”), and compiled into counts per country per lineage per day. The modelling frameworks extends that described in ^20^, and is described and updated at https://github.com/MurrellGroup/lineages with only cosmetic adjustments for the figures shown here.

### Viral isolation, propagation, and titration

All laboratory work involving infectious SARS-CoV-2 occurred under biosafety level 3 (BSL-3) conditions. SARS-CoV-2 variants were isolated and characterised as previously described (EBioMedicine paper). Briefly, diagnostic respiratory specimens that tested positive for SARS-CoV-2 (RT-qPCR, Seegene Allplex SARS-CoV-2) were sterile-filtered through 0.22 µm column-filters at 10,000 x *g* and serially diluted (1:3) on HAT-24 cells (5 × 10^3^ cells/well in 384-well plates). Upon confirmation of cytopathic effect by light microscopy, 300 μL pooled culture supernatant from infected wells (passage 1) were added to Vero-TQ cells in a 6-well plate (0.5 × 10^**6**^ cells/well in 2 mL MEM2%) and incubated for 24-72 h. The supernatant was cleared by centrifugation (2000 x *g* for 5 minutes), frozen at -80°C (passage 2), then thawed and titrated to determine median 50% Tissue Culture Infectious Dose (TCID_50_/mL) on Vero-TQ cells according to the Spearman-Karber method ^21^. Viral stocks used in this study correspond to passage 3 virus, which were generated by infecting VeroE6-TQ cells at MOI=0.025 and incubating for 24 h before collecting, clearing, and freezing the supernatant as above. Sequence identity and integrity were confirmed for both passage 1 and passage 3 virus via whole-genome viral sequencing using Oxford Nanopore technology platform, as previously described^1,22^. For a list of the viral variants used in this study see Supplementary Table S1. Passage 3 stocks were titrated by serial dilution (1:5) in DMEM-5%FBS, mixing with HAT-24 cells live-stained with 5% v/v nuclear dye (Invitrogen, R37605) at 1.6 × 10^**4**^ cells/well in 384-well plates and incubating for 20 h. Whole-well nuclei counts were determined with an IN Cell Analyzer 2500HS high-content microscope and IN Carta analysis software (Cytiva, USA). Data was normalised to generate sigmoidal dose-response curves (average counts for mock-infected controls = 100%, and average counts for highest viral concentration = 0%) and median Virus Effective (VE_50_) values were obtained with GraphPad Prism software. To assess the TMPRSS2 usage of the virus isolates, titrations on HAT-24 cells were performed in the presence of saturating amounts of Nafamostat (20 µM). Titrations were performed in parallel in equivalent volumes of DMSO served as controls and were used to calculate fold drops in VE_50_.

### Abott Architect SARS-CoV2 IgG

IgG antibodies to the nucleocapsid protein (N) of SARS-CoV2 were detected using Architect SARS-CoV-2 IgG (Abbott Diagnostics, Sydney, NSW Australia) as previously described ^23^.

### Rapid high-content SARS-CoV-2 microneutralisation assay with HAT-24 cells (R-20)

Rapid high content neutralisation assay with HAT-24 cells was done as previously described^1,17^. Briefly, human sera or monoclonal antibodies were serially diluted (1:2 series starting at 1:10 for sera) in DMEM-5%FBS and mixed in duplicate with an equal volume of SARS-CoV-2 virus solution standardised at 2xVE_50_. After 1 h of virus–serum coincubation at 37°C, 40 μL were added to an equal volume of nuclear-stained HAT-24 cells pre-plated in 384-well plates as above. Plates were incubated for 20 h before enumerating nuclear counts with a high-content fluorescence microscopy system as indicated above. The % neutralisation was calculated with the formula: %N = (D-(1-Q)) × 100/D as previously described ^2^. Briefly, “Q” is a well’s nuclei count divided by the average count for uninfected controls (defined as having 100% neutralisation) and D = 1-Q for the average count of positive infection controls (defined as having 0% neutralisation). Sigmoidal dose-response curves and IC_50_ values (reciprocal dilution at which 50% neutralisation is achieved) were obtained with GraphPad Prism software. Neutralisation assays testing monoclonal antibodies with VeroE6 cells were performed as previously described^1,17^. Briefly, antibodies were serially diluted in MEM-2%FBS using a 1:3 series starting at 20 µg/mL, input virus solution was standardised at 1.25 × 10^**4**^ TCID_50_/mL, cells were seeded at 5 × 10^**3**^ cells/well in MEM-2%FBS (final MOI = 0.05), plates were incubated for 72 h, and cells were stained with nuclear dye only 2 h before imaging.

### Infection of ALI-pBECs and determination of viral load

Culture and infection of pBEC and alveolar epithelial cultures at air-liquid interface (ALI) was performed as previously described ^24,25^. Briefly, prior to infection, cells were washed once with PBS and inoculated with equal number of virus particles for Delta, Omicron BA.2 and BA.5. Two hours post incubation at 37°C virus inoculum was removed and unbound virus was washed off using 500 μL of PBS. At 3 days post infection cells were collected in RNA lysis buffer (RLT) (Qiagen) with 1% 2-mercaptoethanol for total RNA extraction.

### Statistical analysis

Statistical analyses were performed using GraphPad Prism 9 for all experiments and details have been presented in the figure legends. No statistical methods were used to predetermine sample size. The experiments were not randomised, and the investigators were not blinded to allocation during experiments and assessment of outcomes.

### Data availability

Source data for generating the figures are available in the online version of the paper. Any other data are available on request.

outgrowth

## Results

### SARS CoV-2 Variant diversity through 2022

Through a diagnostic lab-research lab partnership, we isolated SARS-CoV-2 primary isolates throughout 2022. Since the resolution of the BA.1, BA.2 and BA.5 waves, many variants have emerged. However, unlike prior Omicron waves, a single variant is yet to dominate globally. Rather, there exists a pool of variants that have accumulated similar Spike polymorphisms, which can be grouped into sub-lineages derived from BA.2.75 or BA.5 (Fig. 1A-B). Following the peak of the BA.5 wave we focussed on variant isolation and phenotyping to determine the relative risk of newly emerging variants (Supplementary Table 4). All samples were subjected to ex vivo whole genome sequencing using Nanopore single molecule sequencing platforms as previously described^22^. Whilst samples in October were enriched for many BA.5-derived lineages, by mid November there was a similar proportion of variants either derived from a BA.5 parent (e.g. BQ.1) or a BA.2.75 parent (Fig. 1A). The latter was primarily the BR.2.1 variant, with a unique polymorphism within open reading frame (ORF) 8 (S67F). Genomic surveillance supported its origin being within the New South Wales region of Australia (https://www.health.nsw.gov.au/Infectious/covid-19). A snapshot over October to December across Australia (Genomic surveillance through GISAID genome contributions) highlighted the dominance of the recombinant XBF alongside BR.2.1 (Fig. 1B and C). This was alongside the early appearance of other global variants such as XBB.1 and BQ.1.1. (Fig. 1B). By the end of December 2022, XBF represented the dominant variant (Fig. 2C).

**Figure 1.**
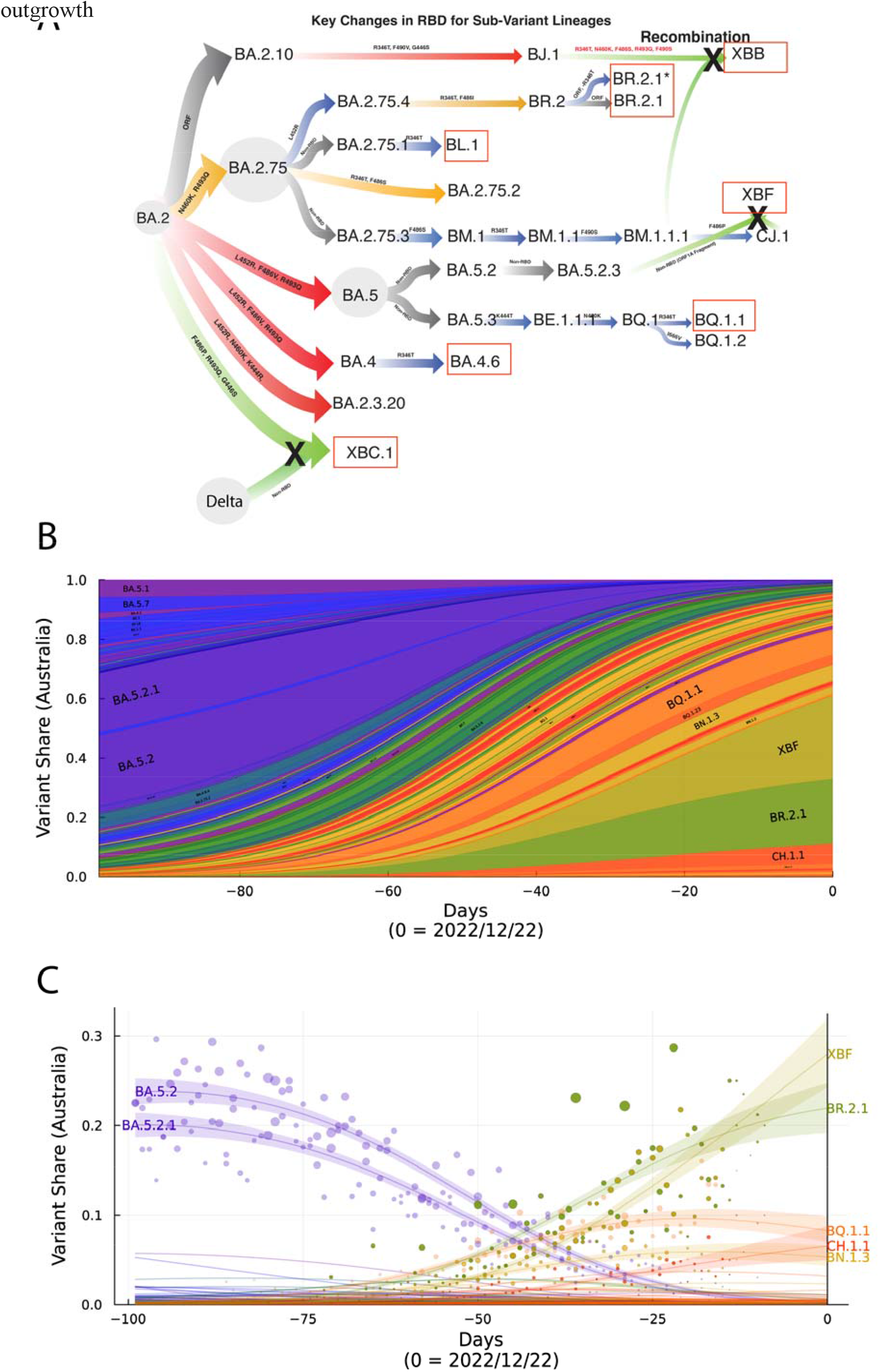
Emergence and prevelance of BA.2.75 and BA.5 sub-lineages with convergent Spike polymorphisms in Australia. **A**. From left to right, Omicron lineages from the parent BA.2 are now primarily split across BA.2.75 and BA.5 sub-lineages. Divergent lineages such as BR.2.1 have accumulated polymorphisms common to BA.5, including L452R and F486I. Whilst BQ.1.1 has acquired BA.2.75 polymorphisms K444T and N460K. Common across most lineages is the additional acquisition of R346T, and this change defines lineages BL.1 and BA.4.6. XXB.1 is a recombinant between BJ.1 and BM.1.1.1 and has benefited from many Spike changes through this recombination event. Whilst XBF is a recombinant between BA.5.2.3 and CJ.1 but retains the latter Spike defined by the F486P change. XBC.1 is a Delta -BA.2 recombinant, with the rention of an intact BA.2 Spike with F486P, G446S and R493Q. Boxed in red are variants that were selected for testing in this study and range from variants with few spike polymorphisms (BL.1 and BA.4.6) through those with many convergent polymorphisms (BR.2.1, XBF and BQ.1.1). For the latter, we have also looked a two circulating isolates of BR.2.1, with and without the R346T change. **B**. Surveillance summary of Australia at 22/12/22 based on genomic data via GISAID. Each vertical slice depicts the posterior mean variant frequency estimate from a hierarchical Bayesian multinomial logistic model of variant competition. Note the two dominant variants BR.2.1 and the recombinant XBF which are dominating in the states of NSW and Victoria respectively. **C**. Variant proportions with associated Bayesian Credible Intervals for **B**.

Whilst isolations were confined to a singular diagnostic unit, statewide genomic surveillance supported similar prevalence of BA.5 and BA.2.75-derived lineages (https://www.health.nsw.gov.au/Infectious/covid-19). Importantly, viral isolation rates were high with over 70% of samples culture positive when samples were restricted to diagnostic Ct thresholds of less than 30. Furthermore, it enabled the curation of isolates that covered both dominant BA.5 and BA.2.75 lineages circulating globally for testing across immunotherapeutics, cohort sera (for antibody evasion) and cell platforms (for viral tropism) (Fig. 2-5). Many of the emerging lineages from divergent parents (BA.2.75 and BA.5) had accumulated polymorphisms clustered and distributed across class I, II and III antibody sites across the receptor binding domain^26^. These included R346T, L452R/M, F486V/I/P, N460K, K444R/T and F490S (Fig. 1), with variants such as XBB.1, BR.2.1 and BQ.1.1 accumulating the greatest number of polymorphisms (Fig. 1).

**Figure 2.**
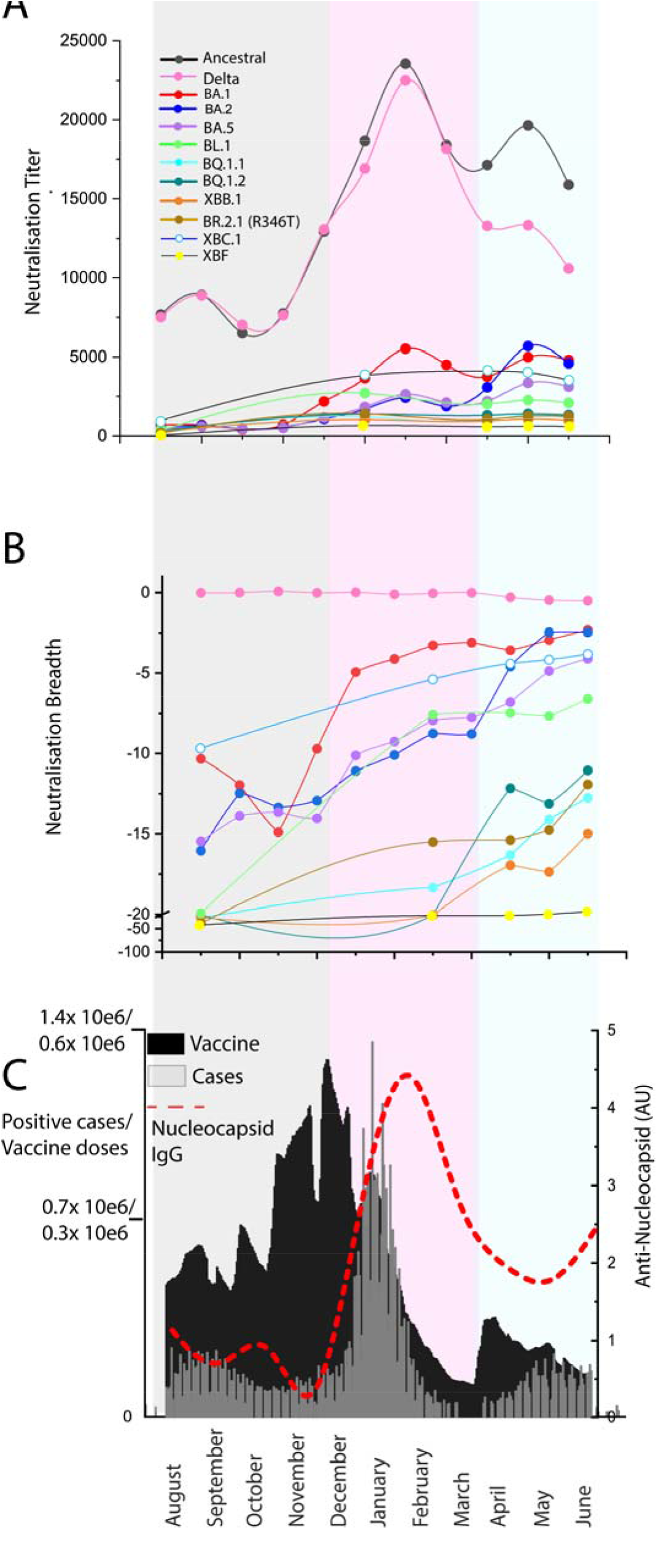
Potency and breadth in pooled IgGs of greater than 420,000 U.S. plasma donors from August 2021 through to June 2022. **A**. Neutralization titers across 12 clinical isolates and include Clade A2.2 (Ancestral), Delta and Omicron BA.1, BA.2, BA.5, BL.1, BQ.1.1, BQ.1.2, XBB.1, XBC.1, XBF and BR2.1 (with R436T) for IgG batches collected between August 2021 and June 2022 (batch details are presented in Supplementary Table I). Each point represents the mean titer of batches from that month and consititutes approximately 30,000 plasma donations. **B**. Breadth score of batches to variants in A. Breadth score is calculated as 1- (fold reduction of titer relative to the Clade A variant A.2.2). When the breadth approaches zero (e.g. Delta), neutralization titers cover this variant with equal potency to the ancestral variant. **C**. Documented vaccines doses and COVID-19 cases are presented here to frame the responses in A & B. Antinucleocapsid IgG levels in monthly batches presented in A. and B. are also indicated (red line). The

### Maturation of viral neutralization breadth over the course of the BA.1 and BA.2 waves and erosion of neutralization titers with emerging Omicron lineages

Herein our primary aim was to track the maturation of the neutralization response over time at the population level. Firstly, we define maturation of the neutralization response as the increasing ability to recognize (increasing neutralization breadth) genetically diverse SARS-CoV-2 variants over time. Secondly, we define neutralization responses at the population level in an approach called IgG pooling. IgG pooling and purification is used in the manufacture of 10% IgG for clinical use in conditions such as replacement therapy for primary and secondary immunodeficiencies. Each batch consists of IgG purified andpooled from in excess of 10,000 plasma donations collected over the period of approximately one month.

To track the maturation of the neutralization response, we analyzed sequential pools of IgG derived plasma from over 420,000 U.S. donors acquired during the period between August 2021 through to June 2022. This time frame covers the acceleration in the 3^rd^ dose (first booster) vaccine roll out in late 2021, the arrival and peak of the largest infection wave in early 2022 (BA.1 Omicron wave) and concludes following the resolution of the Omicron BA.2 wave.

37 separate, sequential batches of pooled IgG, which were tested for neutralization titers across 10 clinical isolates. For variant testing, we look at circulating isolates at that time and also variants that are presently circulating. Isolates included Clade A.2.2 (Ancestral), Delta as well as 2022 Omicron variants, BA.1, BA.2 and BA.5 and emerging Omicron variants BL.1, BQ.1.1, BQ.1.2, XXB.1, XBC.1, XBF and BR.2.1 (Fig. 2A-C). For all polyclonal IgG batches, we tested neutralization titers to each variant (Fig. 2A) and calculated neutralisation breadth to each variant (Fig. 2B). For breadth scoring, scores that approach zero have breadth equal to the “Ancestral” Clade A variant which provides the basis of the vaccines available to the U.S. population (Fig. 2B).

To determine whether the changes in breadth and potency were associated with vaccination versus BA.1 infection, we plotted mean neutralization titers and breadth with three key comparators. Firstly, we plotted the mean titers of batches by month in which the plasma was collected (Fig. 2A). Secondly, we have aligned these titers relative to the breadth to each variant tested. Thirdly, we have aligned documented COVID-19 cases versus vaccine doses in the U.S. population over this time (Fig. 2C) to chronologically frame the immunological events of this population over time. For this, we have divided this time period (Fig. 2C) into the pre-Omicron booster uptake period (grey), Omicron BA.1 wave (pink) and BA.2 wave (blue). To independently track infections, we have also detected anti-nucleocapsid IgG in each batch over time, as neutralization titers associated with either low or high nucleocapsid IgG can independently track and verify if titers may be associated with either vaccine versus infection, respectively (Fig. 2C red dashed line).

The accelerated increase in neutralization titers and breadth to the first Omicron Clade BA.1 (and to a lesser extent BA.2), was associated with the uptake of the third dose of the vaccination regimen. A drop in anti-NC IgG levels relative to neutralization titer, suggested that vaccination, not infection, was contributing to increases in neutralization and breadth during this period (Fig. 2A-C; see grey shading and red line in C). In contrast, we observe a peak of neutralization titers to the Ancestral Clade and anti-NC IgG in February, which was associated with resolution of the BA.1 peak in January 2022 (Fig. 2A-C). The BA.1 wave drove peak neutralization titers to all variants (Fig. 2A-C; see pink shading).

Following the resolution of the BA.1 wave in the U.S., the defining immunological events were a small peak in vaccination doses followed shortly by primarily the BA.2 wave (Fig. 2C). Unlike that observed in early 2022, it was not possible to temporarily dissect vaccination doses from peak COVID-19 cases, and as such, all that can be concluded is that concurrent vaccine uptake and infection within the population led to increasing neutralization breadth to Omicron variants BA.1, BA.2, BA.5, BL.1 and XBC.1 (Fig. 2A-C; blue shading). Across the panel of current circulating variants (BQ.1.1, BQ.1.2, XBB.1, BL.1, BA.4.6 and BR.2.1 (R346T)), the most neutralisation evasive variants clustered together (BQ.1.1, XBB.1, XBF, BR.2.1 (R346T)). In IgG batches from late 2021, there was a 30-50 fold reduction in neutralisation capacity for these variants relative to titers of the Ancestral clade. Whilst this remained largely unchanged, there was an increase in neutralisation breadth to all variants but XBF just prior to the BA.2 wave (Fig. 1A-C; Blue shading). Anti-Nucleocapsid levels remained stable during this period and started to increase following the peak of the BA.2 wave. Thus increasing breadth of neutralisation responses across this period appeared multifactorial but may simply represent the effects of the two dominant immunological events in late 2021-early 2022, i.e. A large proportion of the population receiving their third vaccine dose and/or becoming infected with Omicron BA.1.

### Assessing the relative threat of the late 2022 Omicron sub-variant swarm across a continuum of individual immunity

Whilst testing of pooled IgG from thousands of U.S. plasma donors provides a snapshot of immunity at a population level, cohorts studies where donors have been followed can enable resolution and contributions of vaccination and/or convalescence to the current immune response in the community. As apparent in the pooled IgG approach there is a continuum of immunity ranging from vaccination and/or infection.

To better understand this, we tested the neutralizing response of serum from the Australian Vaccine, Infection and Immunology Collaborative Research cohort (VIIM) with known vaccination and infection histories who donated in late 2022. As time also appears to play a key role in driving viral breadth, we concurrently tested serum from the ADAPT cohort, in which individuals have been followed longitudinally for over two and half years following their first infection in early 2020 and subsequent vaccination. Rather than primarily testing peak vaccine or infection-induced responses, we focused on grouping cohort donors based on their current vaccine and/or infection status. Not only would this provide a snapshot of neutralisation responses at this point in time, but also would enable to determine the relative threat of many emergning variants with convergent polymorphisms. For this analysis, we separated donors into four groups; (i) three dose Pfizer mRNA vaccination with a post-vaccination period of approximately six months (Fig. 3C); (ii) three dose Pfizer mRNA vaccination with breakthrough infections primarily during the overlapping BA.2/BA.5 wave (June to August 2022; Fig. 3A); (iii) four doses of primarily Pfizer vaccine, with the last dose within the last three months of testing (Fig. 3B) and; (iv) donors that were infected during March to August 2020 and subsequently received primarily three vaccinations starting in late 2021 and with a Pfizer vaccine boosters in early 2022 (Fig. 3D). All samples from the VIIM cohort (Fig. 3A-C) were collected in September 2022, as per VIIM vaccine cohort protocol. The ADAPT cohort samples (Fig. 3D) were collected over the period of March to September 2022 and thus within 3 to 6 months post the last vaccine dose. For variant testing we consolidated testing to the group of variants that sustained the greatest fold drops in neutralisation titers relative to the Ancestral variant in pooled IgG studies. This included BQ.1.1, XBB.1, BR.2.1 (+R346T) and also the BA.5.2.3/CJ.1 recombinant XBF which was dominant in Victoria Australia at this time. We also included isolates BL.1 BR.2.1 (346R), BA.4.6, Delta-BA.2 recomginat XBC.1 and BA.5, to determine if limited changes to Spike, such as R346T had significant influence on neutralisation titers at the individual level.

**Figure 3.**
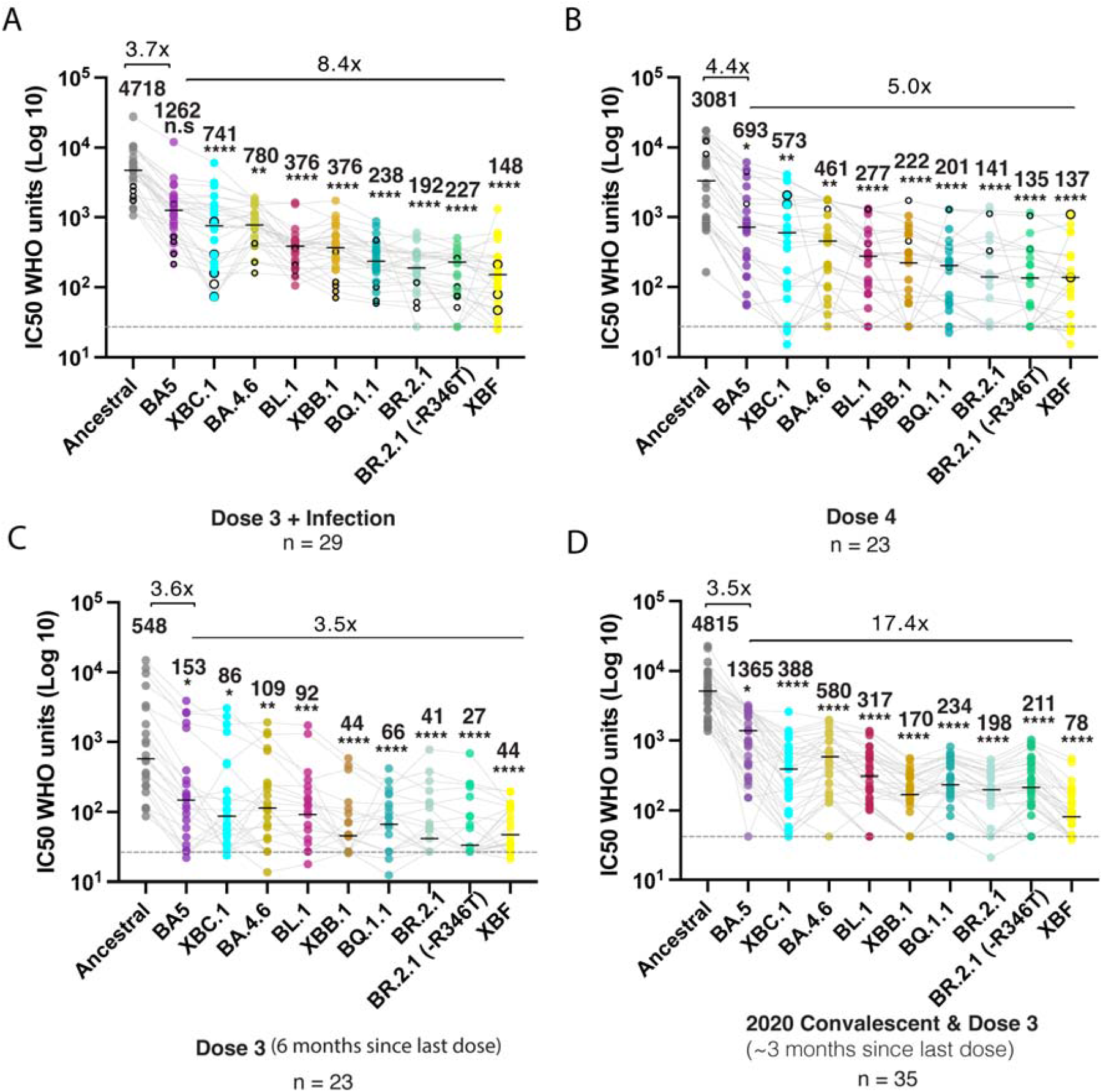
Emerging variants and their ability to evade a continuum of antibody responses. **A**. Three dose Pfizer BNT162b2 vaccination and then a breakthrough infection. Closed circles were infections between June and August 2022 when BA.2 and BA.5 were prevalent. Open circles are breakthrough infections with BA.1 between January and February. **B**. Primarily four dose BNT162b2 vaccinations, in which the last dose was within three months. Open circles in this group are breakthrough infections in early 2022 at the time of BA.1. **C**. Three dose Pfizer BNT162b2 vaccination with the last dose six months prior. **D**. Early 2020 donors infected between March and August of 2020 and then subsequently vaccinated with two doses of Pfizer BNT162b2 or AstraZeneca AZD1222/ChAdOx1 and boosted with Pfizer BNT162b2 or Moderna mRNA-1273. This group did not received their last dose three to six months prior. Data in **(A-D)** indicates the mean IC_50_ of technical replicates for individual samples. The median titers are labelled. Fold change reductions in IC_50_ neutralization titers compare variants of concern to the ancestral variant and Omicron BA.5 where indicated. *p<0.05, **p<0.01, ***p<0.001, ****p<0.0001 for Kruskal Wallis test with Dunn’s multiple comparison test.

BQ.1.1, XBB.1, BR.2.1 and XBF were ranked the most evasive variants across all groups. Whilst three vaccine doses plus infection, four vaccine doses and early convalescent and vaccinated cohorts all observed high titers to the ancestral clade, they showed a 22 to 60 fold drop in titers when tested against XBB.1, BQ.1.1, BR.2.1 XBF (Fig. 3 A. B D). In the cohort with three vaccine doses and a follow up period of 6 months since the last vaccine dose, we did observe significant declines (p <0.001) in neutralisation titers compared to the other three groups. Fortunately, the fold reduction in titers to all variants tested was not at the same magnitiude as that observed across other groups, with only a 12.6 fold drop in neutralisation titers from Ancestral to the most evasive variant tested.

Overall and importantly, similar fold drops and variant rankings were observed using the pooled IgG approach as outlined for US plasma donors and as such supports IgG pooling in monitoring neutralisation responses across large donor collections (i.e. Plasma donation programs and/or national blood banks). If a reduction in neutralization titers will dominate future viral spread, it is likely that multiple contemporary variants will co-exist and persist in the community.

### Emerging variants and resistance to clinical monoclonal antibodies

Clinically utilized monoclonal antibodies (mAbs) such as Evusheld and Sotrovimab are important therapeutics to treat COVID-19 in individuals that have not mounted adequate vaccine responses due to therapy-induced or pre-existing immunodeficiencies. Currently, only Evusheld and Sotrovimab are TGA approved for use in Australia. The neutralising activity of Evusheld (Cilgavimab, Tixagevimab and the Evusheld combination of both) was lost against all variants tested, including BQ.1.1, XBB.1, XBF, BA.2.75.2 and both BR.2.1 lineages (i.e. independent of the R346T polymorphism). Sotrovimab retained neutralization activity against all the above variants except BQ.1.1 and XBF, albeit at a lower potency than A.2.2 (Fig. 4A-B). Given recent studies have observed Sotrovimab neutralisation titers to act as poor indicators of *in vivo* efficacy, we turned to using full length Spike live cell assays ^2^ to sensitively determine if Sotrovimab still bound to variants like BQ.1.1 at levels consistent with that needed for *in vivo* efficacy. As XBF was the last variant tested in our studies, Spike binding data was not available at the time of submission we have focussed primarily on BQ.1.1 herein. Unfortunately spike binding data revealed a 58 fold drop in binding of Sotrovimab to BQ.1.1(Fig. 4C) and thus supports the resistance of this variant in neutralisation assays. In addition, control experiments using XBB.1 and BQ.1.1 spikes with Evusheld further confirmed a lack of spike binding (Fig. 4C) and thus independently confirmed the mechanism for lack of neuralisation observed in these variants.

**Figure 4:**
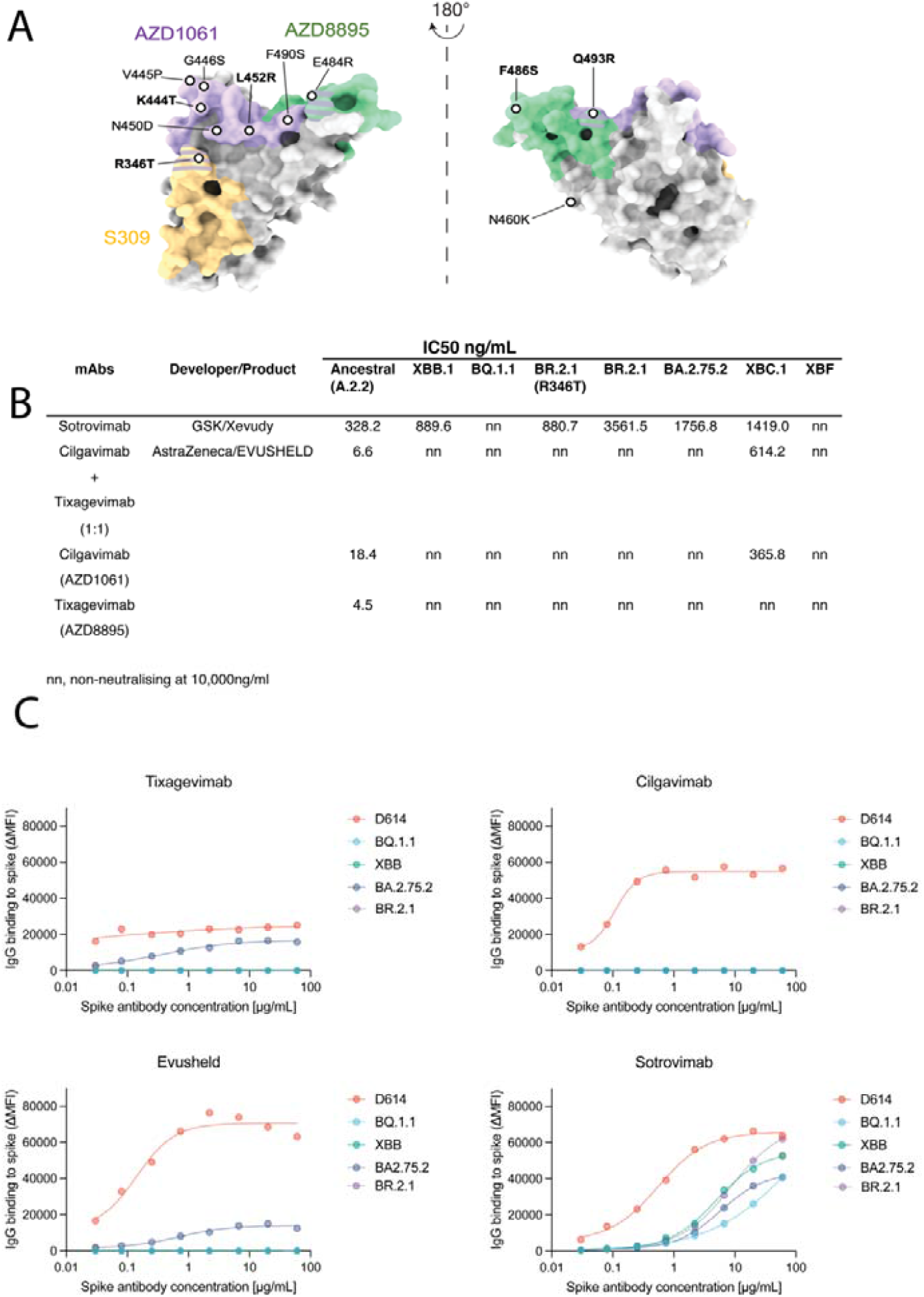
Neutralizing activity of monoclonal antibodies against emerging variants. **A**. Binding sites for AZD1061 (Cilgavimab), AZD8895 (Tixagevimab) and S309 (Sotrovimab) on the SARS-CoV-2 Spike glycoprotein. Key Spike polymorphisms that map to these sites are indicated and are prevelant on emerging variants tested. **B**. IC_50_ values (ng/μl) of monoclonal antibodies Sotrovimab, Cilgavimab and Tixagevimab cocktail, Cilgavimab alone and Tixagevimab alone, against ancestral A.2.2, Omicron XBB.1, BQ.1.1, BR.2.1 (R346T), BR.2.1, BA.2.75.2 and XBF. Antibodies used herein were clinical grade batches. **C**. Antibody binding to full length Spike expressed on live cells and acquired using flow cytometry. Signal is expressed as Mean Fluorescent Intensity above background Fluorescence (ΔMFI).

### Changing TMPRSS2 usage in emerging viral variants

Whilst viral evasion of neutralising antibodies in contemporary samples is a key parameter for assessing potential threat of a variant, other variables can also contribute to increased viral transmission and disease burden in the community. Both ourselves and others have closely monitored changes in viral cell entry requirements and viral cell/tissue tropism associated with emerging variants ^1,7,13,16,17,27,28^. Of particular interest has been the changing use of the serine protease TMPRSS2 in emerging variants. Changes in TMPRSS2 usage can influence viral tropism, which can influence disease severity *in vivo*. For instance whilst Delta and pre-Omicron variants used TMPRSS2 efficiently, the emergence of Omicron variants BA.1 and BA.2 with inefficient TMPRSS2 use was associated changes in viral phenotype. A change with specific preference for epithelial cells derived from the bronchi and the upper respiratory tract was observed ^18^. Whilst disease severity in the clinic has been tempered significantly by vaccination, disease severity in animal models has been correlated with more efficient TMPRSS2 use ^27^.

As changing TMPRSS2 use is key to this change in tropism and potentially an indicator of disease severity, we developed a real-time means of tracking emerging variant TMPRSS2 use by determining endpoint viral titers in a cell line known as HAT-24, that expresses high levels of ACE2 but low levels of TMPRSS2 ^1^. In variants that use TMPRSS2 efficiently, we see a stepped increase in the linear regression established using endpoint titers of primary diagnostic swabs versus their diagnostic qPCR cycle thresholds value (diagnostic Ct or viral load) (Fig. 5A). The hierarchical ranking of variants by infecting HAT-24 cells and determining viral load can readily segregate the pre-Omicron lineage Delta from the early Omicron lineages BA.1 and BA.2 through a downward shift in the linear regression for the latter early Omicron variants. Upon the arrival of BA.5, we observed an increase of TMPRSS2 use above that of its parent BA.2 but lower than that of Delta ^1^. To determine the tropism of this intermediate BA.5 phenotype, we compared Delta, BA.2 and BA.5 infections in primary human bronchial- and alveolar epithelial cultures which were differentiated at the air-liquid interface (ALI). Across both bronchial and alveolar epithelial cell cultures, BA.5 reached viral loads, for a given inoculum, above that of its parent BA.2 and approaching that of Delta after 3 days culture (Fig. 5B). Although after 6 days of culture we observed a significant drop in Omicron lineages BA.2 and BA.5, whilst Delta viral loads persisted at high levels in both primary epithelial cell models (Fig. 5B) Therefore, we conclude the increased use of TMPRSS2 by BA.5 resulted in an early growth advantage across epithelial cells of both the bronchi and lung parenchyma. Furthermore, persistence of high viral loads at day 6 was only observed with the pre-Omicron variant Delta.

**Figure 5.**
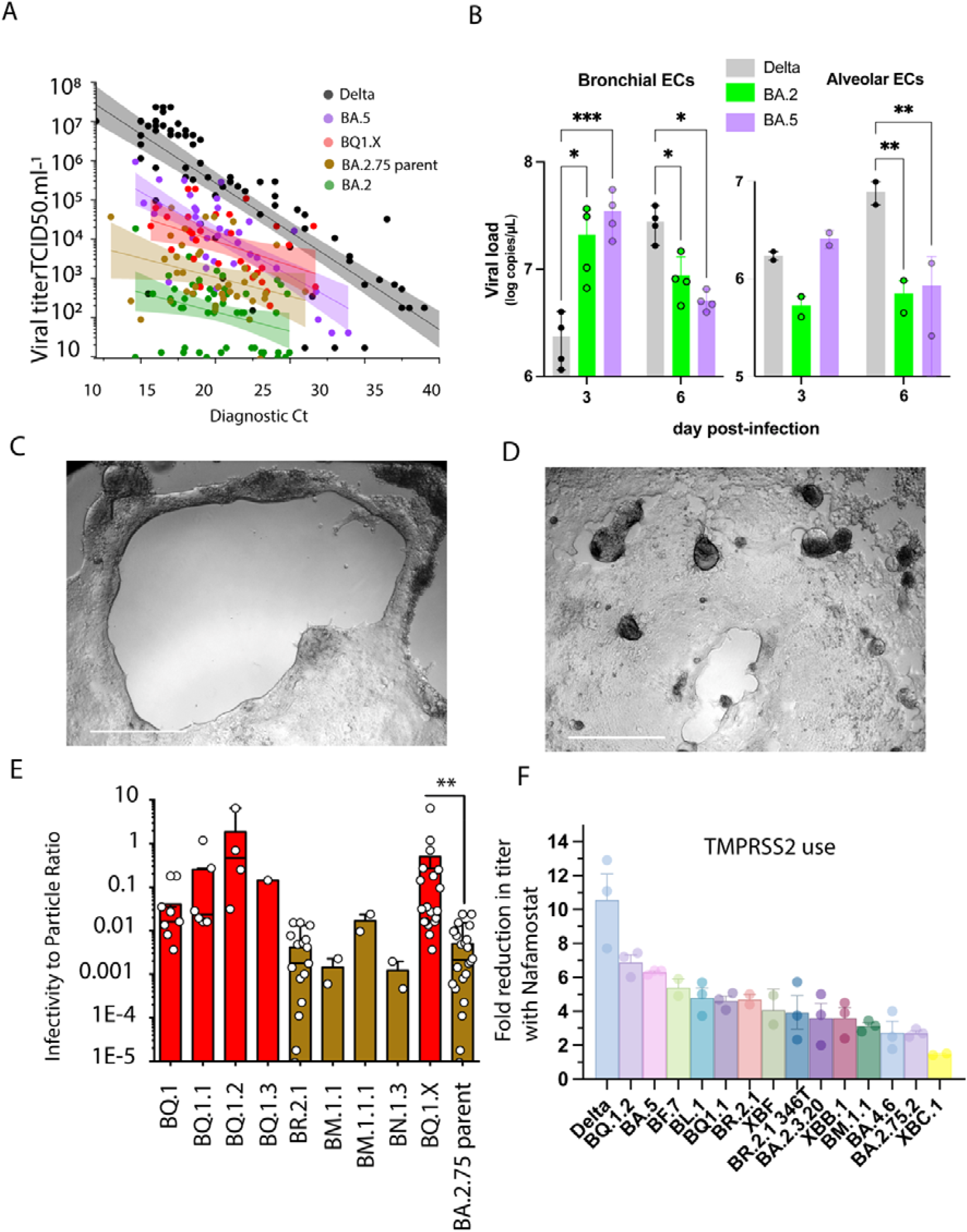
Variant divergence in TMPRSS2 use in the emerging Omicron variant swarm. **A**. Endpoint titers for primary nasopharyngeal swabs grouped into variants derived from BQ (BQ1.X) lineages and variants derived from BA.2.75 lineages (primarily BR.2.1) using the HAT-24 cell line. The linear regressions of BA.2, BA.5 and Delta are also presented ^1^ and serves here primarily as a guide with respect viral titers per diagnostic Ct value. Of note, as the variant uses TMPRSS2 more efficiently, there is an increase in viral titer per Ct value and this is observed by an upwards shift in the linear regression. **B**. Air layer interface (ALI) primary bronchial and alveolar epithelial cells 3 & 5 days post-infection. Equal numbers of viral particles were used to inoculate each culture. Supernatant was harvested 3 & 6 days post-infection. Error bars represent standard deviations from the mean of infections from two to four independent donors. Delta and the parent variants BA.2 and BA.5 are presented here to representative highlight the influence of TMPRSS2 usage for viral entry fitness in primary differentiated respiratory epithelia in the upper and lower respiratory tract. *p<0.01, **p<0.01 ***p<0.001 for two way ANOVA with Sidak’s multiple comparisons **C**. BQ.1.2 and **D**. BR.2.1 primary nasopharyngeal swabs are presented in HAT-24 cells following 3 days in culture. Scale bar represents 100 µm. Limiting viral dilutions are presented to contrast the growth of each variant. **E**. To enable initial resolution of variants, we determined RNA copies per diagnostic Ct values and have presented infectivity to particle ratios for variants that were initially group in A. Grouping of BA.5 and BA.2.75 lineages here highlight the latter group to have lower infectivity to particle ratios. **p<0.01 for Kruskal Wallis test with Dunn’s multiple comparison test. **F**. Mean fold reductions in viral titers in the presence of saturating levels of the TMPRSS2 inhibitor Nafamostat. Mean values represent three independent experiments and each point is the mean fold reduction per independent experiment performed in quadruplicate.

As BA.5 emerged with an outgrowth advantage globally, we hypothesised that both antibody evasion and better TMPRSS2 use was key in outcompeting other variants. To monitor the TMPRSS2 entry phenotype of emerging variants beyond antibody evasion alone, we worked closely with local diagnostic networks to determine endpoint titers of 188 primary swabs (Supplementary Table 4) over the months of October and November 2022. In parallel, we performed whole genome sequencing to link each swab and its endpoint titer to emerging variants. Unlike other waves that we have characterized in the Australian community, we did not observe a dominant variant, but rather the emergence of many genetically distinct sub-variants stemming from BA.5 and BA.2.75 parent lineages (Supplementary Table 4). To resolve this further, we pooled each variant and then determined infectivity to particle ratios based on diagnostic Ct values and endpoint titers. Whilst the sample number of each variant could not lead to statistical testing between variants, the pooled data from BQ variants demonstrated higher infectivity to particle ratios than the pooled variants derived from BA.2.75 (Fig. 5E) using the HAT-24 cell line. Futhernore, monitoring the growth of variants in culture visually demonstrated greater fusogenicity in BQ variants (Fig. 5C) versus BA.2.75-derived variants like BR.2.1 or BA.2.75.2 (Fig. 5D).

Given the diversity of variant sub-lineages restricted the above approach, we resolved the continuum of TMPRSS2 usage across Omicron sub-variants in expanded isolates. In this approach, viral isolstes are titrated on the HAT-24 cell cell line with excess (determined through dose-dependent titrations with the Delta variant) serine protease inhibitor Nafamostat. When a variant (e.g. Delta) efficiently uses TMPRSS2, the presence of excess Nafamostat significantly reduces viral titers (Fig. 5F). In contrast, BA.1 and BA.2 utilized TMPRSS2 poorly and, in turn, this resulted in lower reductions in titers in the presence of Nafamostat. Using this approach, we tested 14 emerging variants that are representative of the present variant swarm. Relative to BA.5, all variants with the exception of BQ.1.2 demonstrated similar or reduced reliance on TMPRSS2 use based on viral titers observed in the presence of Nafamostat (Fig. 5F). As observed in primary nasopharyngeal swabs, there was a trend for BA.2.75 sub-lineages to utilise TMPRSS2 approaching that of BA.2. Of Interest, the Delta-BA.2 recombinant XBC.1 was observed to have the lowest TMPRSS2 use, and this is consistent with the majority of the BA.2 Spike (including RBD and furin cleavge site) being primarily retained in this recombinant. To conclude, using primary swabs and expanded primary isolates, whilst all circulating isolates were engaging TMPRSS2 at levels higher than BA.2, they were at or lower than that observed for BA.5 and importantly on a trajectory to reach that of Delta.

## Discussion

Continued genomic and phenotypic surveillance of emerging variants is of importance as communities learn to navigate the complexities of SARS-CoV-2 viral spread. Antibody responses have been key in understanding and predicting vaccine efficacy, and combined with observations of viral fitness and tropism, can be highly predictive in determining the relative threat of an emerging variant. Herein we observe surprising breadth and potency in the population antibody response driven by exposure to pre-omicron variant spike proteins delivered with by vaccination an/or infection. While this pre-exposure breadth of antibody responses provides a positive observation with regards viral control and protection from disease progression, we also observe the accumulation and convergence of a variant pool within the community which all share various combinations of common Spike polymorphisms. This affords these variants the ability to somewhat better navigate the current broad and often potent antibody responses. Concurrently, we also observe incremental improvements in antibody breadth across recent cohort samples and at the population level using pooled IgGs. It is unclear whether breadth will further increase with the contribution of variant vaccines or post infection with these variants. With respect to viral entry, whilst better utilization of TMPRSS2 use can enable variant outgrowth advantage and expanded tropism to epithelial cells of the lower respiratory tract, we fortunately observed a continuum of TMPRSS2 use towards that of BA.2 as opposed to the increased efficency of TMPRSS2 use observed with pre-Omicron variants like Delta ^1^.

The pooling of IgG from large numbers of plasma donations enabled an unprecedented snapshot of a population’s response to vaccination and the largest COVID-19 wave, Omicron BA.1. For this approach, this study focused primarily on the U.S. population, which benefited from the early roll out and uptake of many mRNA- and vector based vaccine doses in early 2021, with doses peaking in April 2021. This early roll out of vaccines was followed by substantial booster uptake in late 2021, just prior to the U.S. Omicron BA.1 wave. Booster uptake during this period provided a signal of increasing neutralisation titers in the community whilst accelerating breadth of these responses to boost reponses to variants that had yet to enter the community. Whilst the BA.1 wave continued the upward trajectory of neutralisation titers, it was not accompanied with increasing breadth to Omicron BA.1 or BA.2 lineages. Though it is tempting to conclude that increasing breadth is only associated with vaccine booster uptake, it may simply reflect the finite breadth in the community at that juncture of time that cannot increase until further maturation of the immune response can proceed. Increasing neutralisation titers and breadth of mRNA vaccine responses is observed to be relative to that of pre-existing antibody levels, with greater fold increases in antibody potency and breadth observed in those individuals with lower circulating antibodies prior to their vaccine booster dose ^29^. Thus, greater potency and breadth within the community following an additional vaccine dose and/or infection will be limited with either immune encounter. Fortunately over 2022, further monitoriting of pooled IgGs throughout 2022 demonstrated continued increased in variant breadth. Whilst this was primarily against BA.1, BA.2 and BA.5, increasing breadth to emerging variants XBB.1, BQ.1.1 and BR.2.1 was also observed. As was the case prior to the BA.1 wave, vaccination and/or infection can contribute to increased antibody breadth even when a variant is not yet in circulation.

Whilst the use of IgG pooling can give a large population snapshot of antibody-based immunity within the community at a point in time, this approach does not allow resolution of responses at the individual level. Yet in observing many current cohorts, the unifying feature of all responses is greater breadth to the past evasive Omicron variants such as BA.1, BA.2 and BA.5 as well as those emerging within the community. Whilst the waning of antibody responses is of concern and probably important in managing future waves with various vaccine boosters, we observed increased breadth across all Omicron variants even within groups with waning responses (six months post-three mRNA vaccine doses). This is consistent with earlier mRNA vaccine studies, in which neutralizing antibody levels post-second dose stabilized between six and nine months post-vaccination but importantly for the pool of antibodies that persisted, there was continued improvement in their neutralization capacity and breadth over this period ^29^. This is also consistent with our observations in 3 vaccine dose donors where the response is 6 months after the last dose. In this setting, the antibodies that persisted had a greater capacity to neutralise all emerging variants.

Whilst time appears key in greater antibody breadth to the virus, the emergence of the present pool of variants with convergent Spike polymorphisms appears to be the counter response to the immune challenge. In the emerging variants XBB.1, BQ.1.1, BR.2.1 and XBF, we observed significant increases in neutralisation evasion despite the developing breadth of the maturing antibody response across all groups. Of interest is that all three variants have converged to cluster within the same level of evasiveness but with emergence from genetically diverse parental lineages. XBB.1 derived from a BA.2.10 with BA.2.75 lineages recombination event, XBF a BA.5.2.3 recombination with CJ.1 (with the latter spike intact) and BR.2.1 and BQ.1.1 derived from BA.2.75 and BA.5 lineages respectfully. Of obvious concern is the continuing decay of clinical options for those individuals with a limited antibody response following vaccination and/or infection. The lack of neutralization activity of Evusheld across many emerging variants will need to be considered carefully moving forward with respect to clinical treatment. Whilst Sotrovimab maintained activity across many variants, it lost neutralisation activity against BQ.1.1 and XBF, with testing of binding to spike observed to be significantly reduced for BQ.1.1. Whilst the correlates of the *in vivo* activity for Sotrovimab are complex ^30^, treatment in the era of a variant mix will provide many challenges and will require continued monitoring. In the latter treatment setting, the continued discovery of monoclonal antibodies and/or combinations thereof is still warranted, but require combinations of antibodies with inherent breadth. In addition, as the community gains greater potency and breadth of antibodies, revisiting the use of polyclonal antibodies ^31^ harvested from plasma donors may also provide a treatment contingency for those at risk. Taken together these data suggest that boosting of vaccine respnses in immunocompetent individuals should be encouraged as an effect means of boosting effectively antibody responses.

The trajectory of each variant in each global jurisdiction is being monitored carefully at many levels. Variants like XBB.1 have already shown truncation of case waves in Singapore (https://www.moh.gov.sg/covid-19/statistics) and XBB.1.5 is starting to supplant BQ and BA.5 variants the US (www.ecdc.europa.eu/en/covid-19/country-overviews; www.cdc.gov/coronavirus/2019-ncov/variants/variant-classifications.html; covariants.org/per-country). In this setting, Australia is unique, as the global pool of key variants is presently circulating within the community, alongside those which appear to have been seeded locally (e.g. BR.2.1 and XBF). Whilst all variants have converged to share similar Spike polymorphisms that enable antibody evasion, it is presently unclear if this alone will enable an outgrowth advantage in any one variant to drive similar case waves as that seen in prior Omicron waves. XBF for instance is now the dominant variant in Australia and shares a spike profile very similar to that of XBB.1.5 with defining spike polymorphisms such as F486P & F490S, with the former linked to greater affinity for ACE2 binding. Key to understanding the potential of the current convergent pool is whether gaining the ability to evade antibodies has sustained a fitness cost in entering cells/tissue. In this setting, we do observe the overall trend of emerging variants to utilize key entry proteases such as TMPRSS2 less efficiently and lower than BA.5. Moving forward, there will be sustained focus on understanding how this presents in the community across the diversity of global pandemic experiences. Whilst changes to the Spike to avoid antibodies can impact upon viral cell-entry fitness into various tissues, additional changes that may compensate for this fitness loss will need to be tracked carefully. For instance, the accumulation of polymorphisms at and around the furin cleavage site, such as I666V in BQ.1.2 may have limited impact in influencing antibody binding, but rather increase viral cell entry fitness and in turn compensate for any fitness loss in cell entry. Whilst viral entry changes are often associated with increased transmission, it is also associated with expansion or contraction of viral tropism and the former has been observed with signals of increased disease severity not only in animal models^27^ but also in large clinical studies on the recent BA.5 wave^32^. Whilst there are many currently circulating variants, variants that maintain entry fitness and evade antibodies concurrently will have a greater outgrowth advantage to supplant the others and/or result in greater levels of disease severity as a consequence of either increased viral loads and/or changing viral tropism to enable infection of the lower respiratory tract. Although viral entry can primarily influence the latter, evasion of innate immunity through the action of non-Spike viral gene products needs to be also monitored carefully changes thereof may enable high viral load persistence in the respiratory tract and elsewhere to augment events that lead to greater clinical disease severity. Whilst a variant with genomic profile with potential of high disease severity may not arise, continued genomic surveillance and rapid phenotyping as described herein, will be key to identifying any such future variants. Whilst a limitation of this study maybe that *in vitro* data does not necessarily correlate with clinical outcomes, it is imperative that datasets herein and elsewhere are continually assessed alongside clinical observations of therapeutic and vaccine efficacy and presentation of disease severity.

## Data Availability

Source data for generating the figures are available in the online version of the paper in the supplmentary tables. Any other data are available on request

